# Translation, cross-cultural adaptation, and psychometric evaluation of the colour blind quality of life scale into Vietnamese

**DOI:** 10.64898/2026.05.15.26353359

**Authors:** Tran Thi Thuy, Pui Juan Woi, Mohd Izzuddin Hairol, Que Anh Vu

**Author notes:** Corresponding author (PJW).

## Abstract

Background: The Colour Blind Quality of Life Scale (CBQoL) is a questionnaire developed to assess the quality of life of individuals with congenital colour vision deficiency (CVD). This study aimed to translate the English version of the CBQoL into Vietnamese and evaluate the validity and reliability of the Vietnamese version (CBQoL-VN). Methods: A forward-backward translation method was performed to produce the Vietnamese text. Content validity was assessed by six experts in vision care. Reliability testing involved 30 participants with congenital CVD, while discriminant validity was evaluated by comparing this group against 30 participants with normal colour vision. Results: Following expert consensus, two items were removed and one transportation-related item was added. The content validation index (CVI) values of 1.0 for relevance, clarity, and understandability indicated excellent content validity. Internal consistency was high, with a Cronbach’s alpha of 0.95 for the full scale. Discriminant validity analysis showed that participants with congenital CVD scored significantly lower across all CBQoL-VN domains compared to those with normal colour vision. Conclusions: The modified CBQoL-VN is a valid and reliable instrument for assessing the quality of life of individuals with congenital CVD in the Vietnamese population.

## Introduction

Congenital colour vision deficiency (CVD) is a common hereditary visual anomaly resulting from mutations in cone photopigment genes located on the X chromosome [1]. It affects approximately 8% of men and 0.4% of women worldwide [2], with variations by ethnicity and geography. While often regarded as a mild disability, congenital CVD can impose significant challenges on an individual’s daily life, educational performance, and career opportunities [3,4]. Individuals with CVD often face difficulties in tasks that rely on colour discrimination, such as interpreting maps, identifying ripe fruits, and distinguishing traffic signals.

Beyond functional limitations, CVD can have profound psychosocial impacts. Research indicates that individuals with CVD may experience embarrassment, anxiety, and a sense of inadequacy due to their inability to perform colour-related tasks correctly [5–7]. Furthermore, occupational restrictions in fields such as aviation, law enforcement, and engineering can lead to frustration and limited career progression [8]. Despite these impacts, clinical assessments typically rely on diagnostic tools like the Ishihara plates or the Farnsworth D-15 test. While these tools effectively diagnose the presence and type of deficiency, they fail to capture the subjective burden of the condition on the patient’s quality of life.

To address this gap, Barry et al. (2017) developed the Colour Blind Quality of Life Scale (CBQoL), a valid and reliable instrument designed specifically to measure the impact of CVD across health, emotional, and occupational domains. However, questionnaires are culturally and linguistically sensitive; a direct translation is often insufficient to ensure an instrument is valid for a different population [9]. Differences in lifestyle, environmental factors, and cultural norms necessitate a rigorous process of cross-cultural adaptation to ensure conceptual equivalence.

Currently, there is no validated instrument in Vietnam to assess the quality of life for individuals with congenital CVD. Simply translating an existing Western tool is insufficient due to Vietnam’s distinct environmental and lifestyle characteristics. Regarding transportation, the country is characterized by a high-density, mixed-traffic system dominated by motorcycles [10]. According to the 2024 Population and Housing Survey by the National Statistics Office of Vietnam (2024), only 9% of Vietnamese households own cars, whereas 89.4% utilize motorcycles. Unlike the highly structured road systems in Western countries, navigating Vietnamese traffics requires rapid visual discrimination of signals and brake lights within a chaotic traffic flow. These unique conditions pose specific safety risks and functional challenges for individuals with CVD that may not be captured by international instruments.

Furthermore, daily living in Vietnam heavily involves traditional open markets, where the selection of fresh products, such as distinguishing ripe fruit or fresh meat based on subtle colour cues is a central daily activity. These specific environmental demands differ significantly from the context in which the original CBQoL was developed. Consequently, a specific adaptation of the scale is required to accurately capture the unique visual challenges and safety concerns faced by the Vietnamese population. Therefore, this study aimed to (i) translate and cross-culturally adapt the original CBQoL into Vietnamese (CBQoL-VN) and (ii) evaluate its validity and reliability within the Vietnamese population.

## Materials and methods

### Study design

This study had two phases: (i) Phase I: translation of the English version of the CBQoL into Vietnamese language and (ii) Phase II: validation and reliability testing of the CBQoL-VN. The stages of the translation and validation process are summarized in Fig 1. This study was approved by the Research Ethics Committee of Universiti Kebangsaan Malaysia (JEP-2025-061).

**Fig 1.**
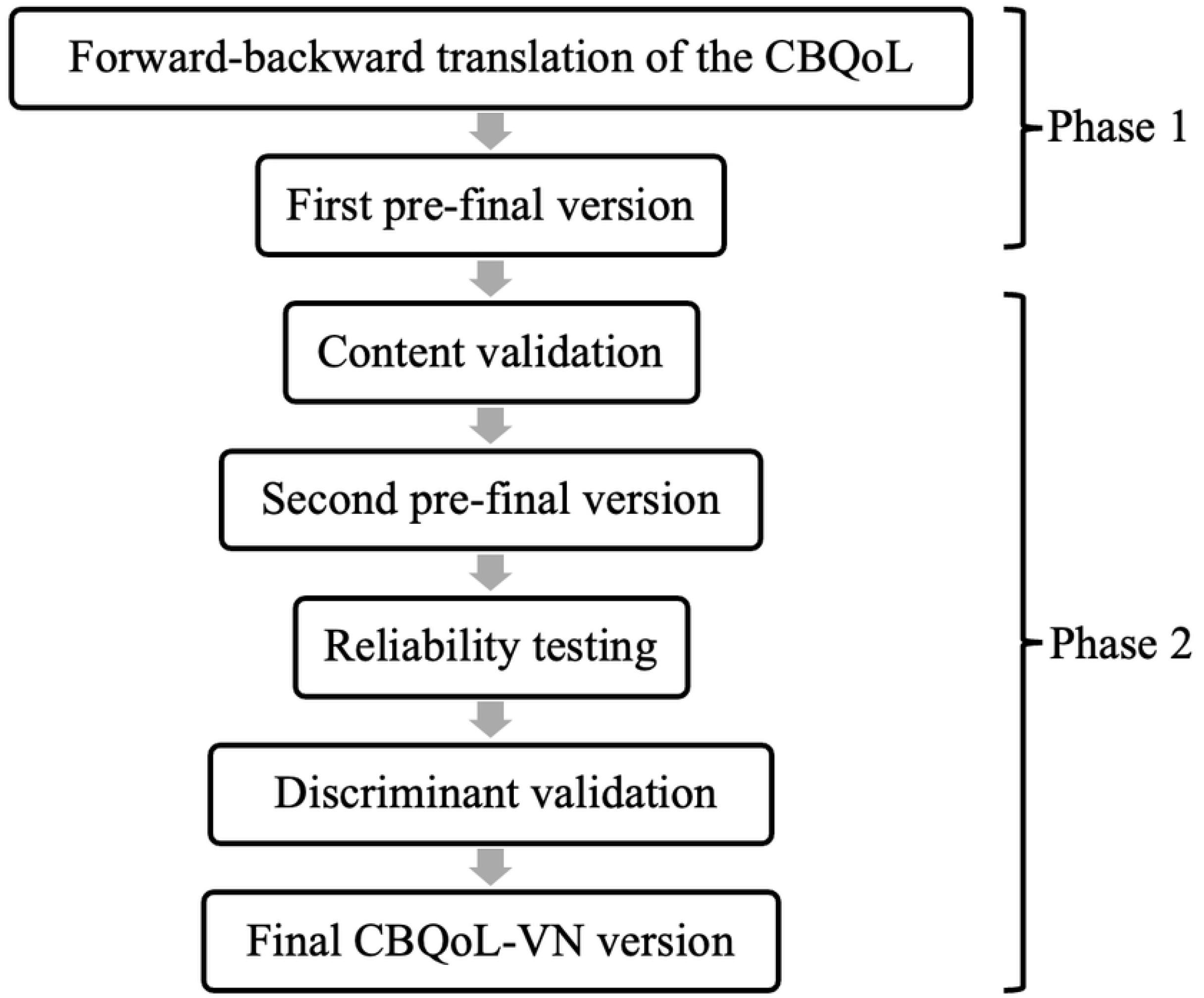
Flowchart of the CBQoL-VN translation and validation process.

All procedures were conducted in accordance with the Declaration of Helsinki. Recruitment of this study started on 21 February 2025 and ended on 31 March 2026. Electronic or printed informed consent was obtained from all experts and participants in the study.

### Instrument

The open-access original English version of the CBQoL is useful to evaluate the quality of life in relation to health, emotional life, and work in individuals with congenital CVD [7]. It is a 23-item self-reported measure of quality of life that uses a six-point Likert-type scale format, with responses ranging from 1 = “A severe problem” to 6 = “No problem”, with an option for “not applicable” (N/A) (Table 1). Each domain score is calculated as a mean, ranging from 1 (minimum) to 6 (maximum). If a response was marked as ‘N/A’, the mean of the remaining items is used, provided that at least 66% of the participant’s responses within that domain are valid numeric (non-N/A) values.

**Table 1.**
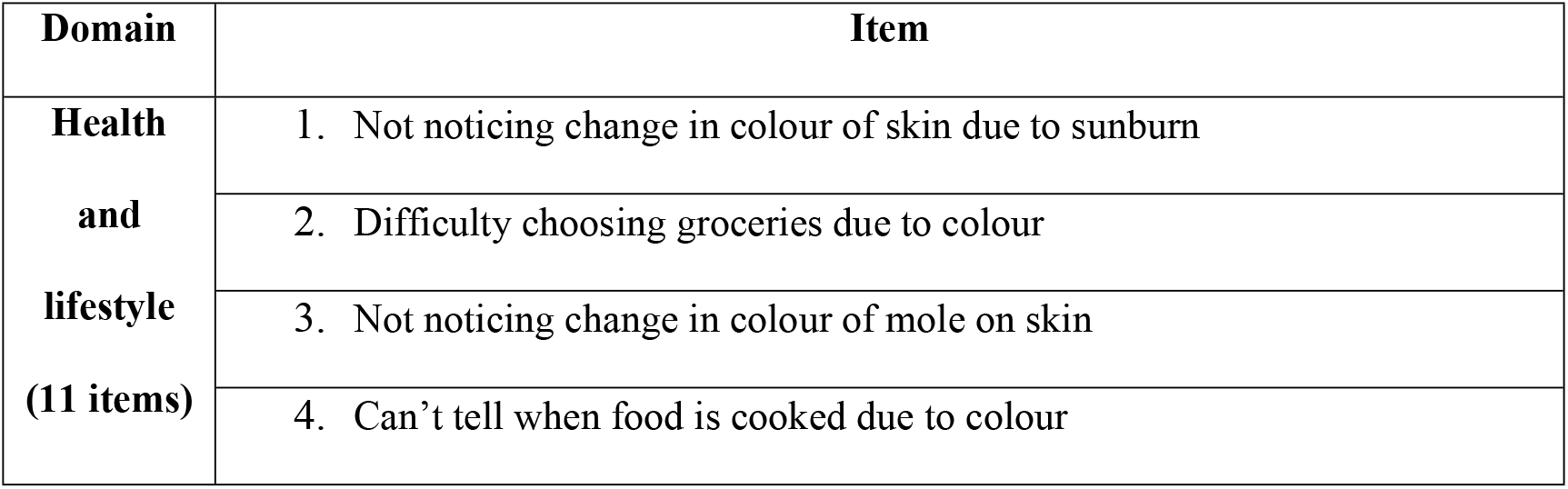
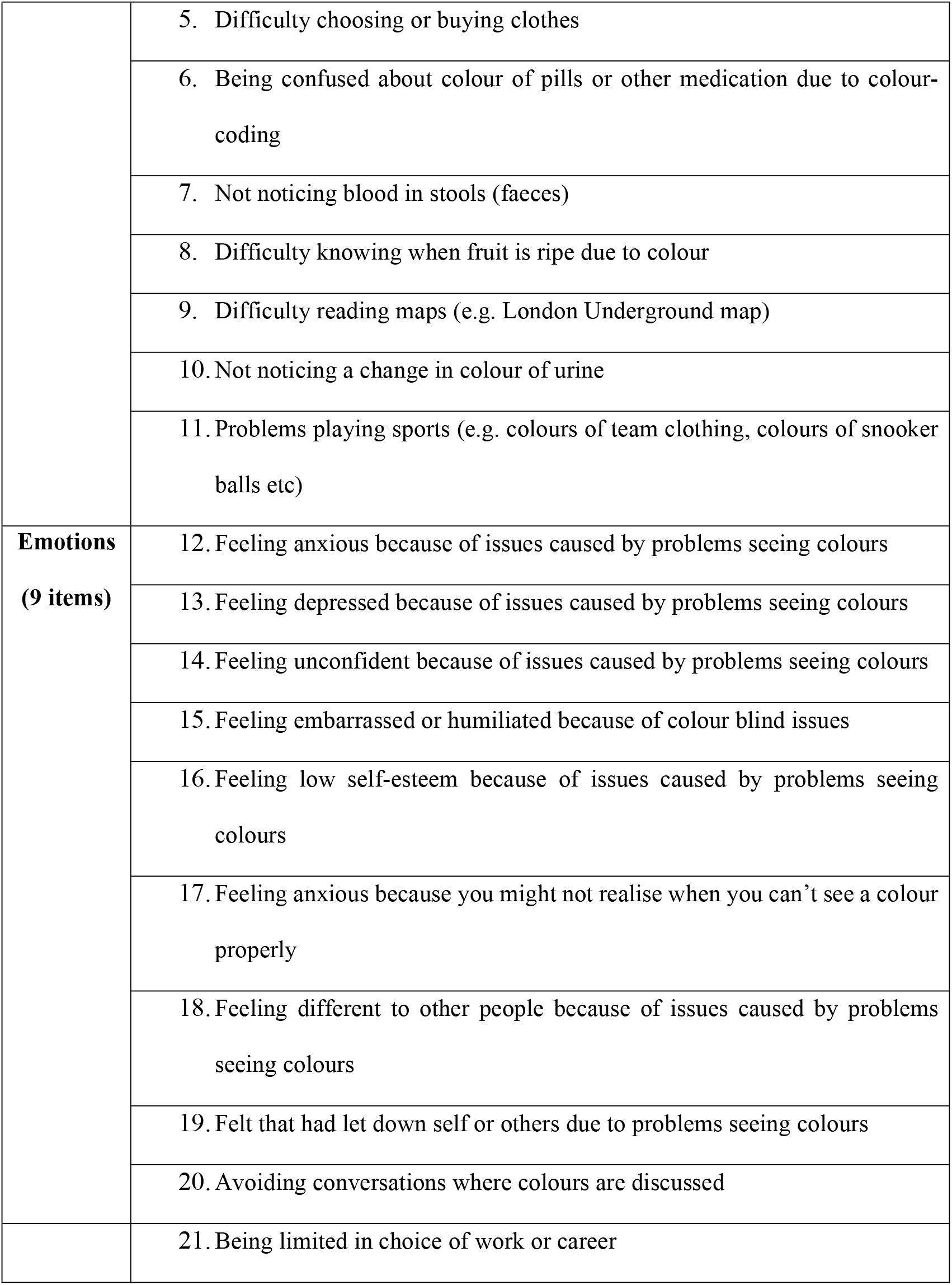
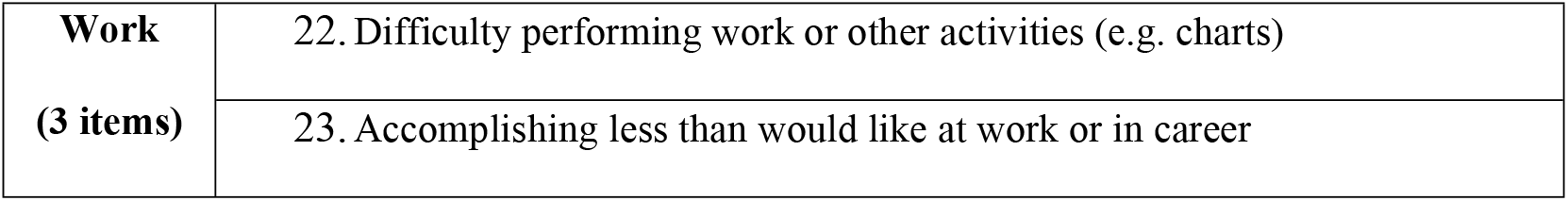
The original English version of the CBQoL.

### Phase I: Translation

Translation was performed according to the World Health Organization’s (WHO) guidelines on translation and adaptation of instruments, which included forward and backward translations. The English version of the CBQoL was first translated into Vietnamese language with forward translation by a bilingual subject matter expert. The CBQoL-VN was then translated into English using backward translation by a linguistic expert with experience in the medical field. To produce the first pre-final version, both the original and the backward translated English versions of the CBQoL were reviewed by another linguistic expert, who compared them and checked for discrepancies.

### Phase II: Validation and reliability testing

#### Content validation

Six bilingual experts in eye health were selected for the content validation of the CBQoL-VN. The expert group comprised three ophthalmologists, two clinical optometrists from academic institutions, and one optometrist from a primary care practice. All experts received written information via email about the objective of the study and their role as expert panels.

In the first round, the experts assessed all items in the CBQoL-VN for relevance, clarity, and comprehensibility, providing written comments for each item. A discussion was conducted among them to identify and resolve the inadequate expressions or concepts of the translation, as well as any discrepancies between the Vietnamese version and the original English version of the CBQoL. All feedback was carefully considered and used to refine the questionnaire, resulting in an improvised version. This second pre-final version of the CBQoL-VN was then redistributed to the experts for further evaluation. In this round, they rated each item using a 4-point Likert scale, where 1 indicated the lowest and 4 the highest value. All experts completed and returned the questionnaires within two weeks.

#### Reliability

A total of 30 participants (mean age = 26.70 ± 9.34 years) were recruited through purposive sampling for the reliability testing of the modified CBQoL-VN following content validation. Participants met the following inclusion criteria: Vietnamese nationality, proficiency in Vietnamese, age between 19 and 60 years, and a confirmed diagnosis of congenital CVD. The diagnosis of congenital CVD was confirmed using Ishihara plates (38-plate) and family history.

All participants completed the questionnaire via an openly accessible online survey platform, Google Form or printed form. The questionnaire consisted of two parts: demographic data and the modified CBQoL-VN. The data were gathered over a 6-month period. The internal consistency of the modified CBQoL-VN was assessed using Cronbach’s Alpha. Table 2 shows a summary of the participants’ demographic characteristic.

**Table 2.**
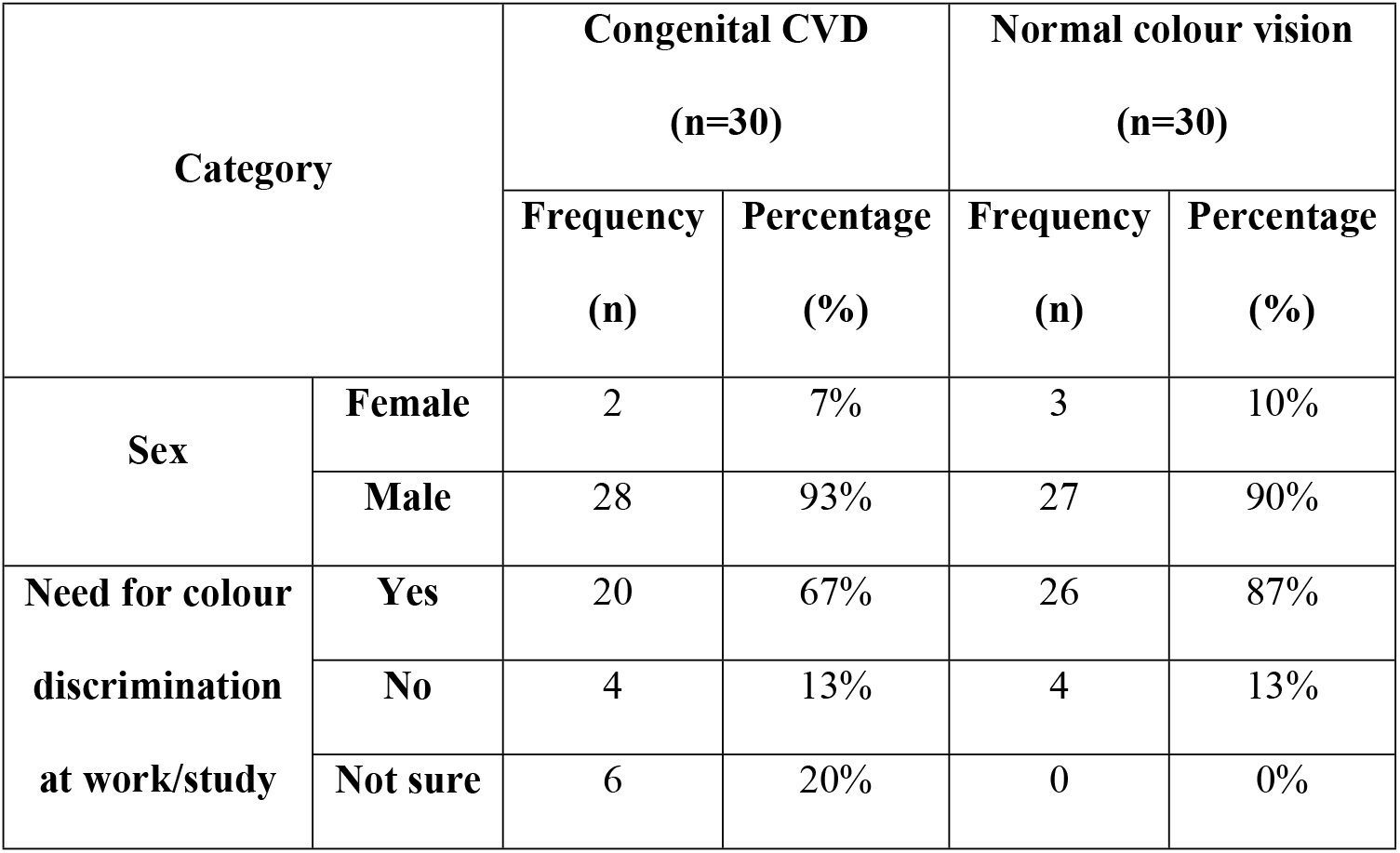
Demographic characteristic of participants (N=60).

| Category |  | Congenital CVD<br>(n=30) |  | Normal colour vision<br>(n=30) |  |
| --- | --- | --- | --- | --- | --- |
|  |  | Frequency | Percentage | Frequency | Percentage |
|  |  | (n) | (%) | (n) | (%) |
| Sex | Female | 2 | 7% | 3 | 10% |
|  | Male | 28 | 93% | 27 | 90% |
| Need for colour discrimination at work/study | Yes | 20 | 67% | 26 | 87% |
|  | No | 4 | 13% | 4 | 13% |
|  | Not sure | 6 | 20% | 0 | 0% |

#### Discriminant validity

A comparison group of individuals with normal colour vision (mean age = 23.47 ± 2.30 years) was recruited for discriminant validity testing. Discriminant validity was evaluated using a known-groups comparison approach, in which participants with congenital colour vision deficiency (CVD) were compared with those with normal colour vision. Total CBQoL-VN scores and domain scores (health and lifestyle, emotions, and work and study) were calculated for each participant.

### Statistical analysis

The content validation index (CVI) was calculated based on the experts’ ratings. Two types of CVI were calculated in this study, which were item-level content validity index (I-CVI) and scale-level content validity index (S-CVI) [12]. Two methods were performed to calculate S-CVI, which were the average of the I-CVI scores for all items on the scale (S-CVI/Ave) and the proportion of items on the scale that achieve a scale of 3 or 4 by all experts (S-CVI/UA) [13]. The S-CVI/UA was based on the universal agreement (UA) method, where 1 was given when the item achieved 100% agreement from experts, otherwise the UA score was given as 0. The acceptable CVI value was at least 0.83 for an expert group of six [13,14].

The Statistical Package for Social Sciences (SPSS) Version 28.0 (IBM Corp, Armonk, NY) was used to perform reliability analysis. An alpha coefficient greater than 0.7 was considered to have good reliability [15]. The normality of data was tested with the Shapiro-Wilk test. All data sets were not normally distributed. The discriminant validity was evaluated by comparing total CBQoL-VN scores between congenital CVD and normal groups using Mann-Whitney U test. The p-value was 2-sided, and a statistical level of p < 0.05 was considered significant.

## Results

### Content validation

In the first round of review, five out of six experts noted that some items conveyed overlapping meanings when translated into Vietnamese. Specifically, Item 12 (“Feeling anxious because of issues caused by problems seeing colours”) was considered similar to Item 17 (“Feeling anxious because you might not realise when you can’t see a colour properly”), and Item 14 (“Feeling unconfident because of issues caused by problems seeing colours”) was similar to Item 16 (“Feeling low self-esteem because of issues caused by problems seeing colours”). After discussion, the experts reached consensus to remove Items 16 and 17 from the emotional domain.

The panel also discussed potential redundancy between Item 2 (“Difficulty choosing groceries due to colour”) and Item 8 (“Difficulty knowing when fruit is ripe due to colour”). To avoid overlap, Item 8 was revised to include additional terms such as flowers and food, thereby distinguishing it from groceries. In this context, “groceries” refers to items other than fruits, flowers, or food. Items 3 and 22 were rephrased to enhance clarity, while Item 9 was revised to better fit the context of Vietnamese daily life.

Ultimately, no further items were removed, but one transportation-related item (Item 12) was added to the health & lifestyle domain. Following the discussion and consideration of all expert feedback, the order of items within each domain was rearranged to facilitate participant recall. Table 3 shows the 22 items of the modified CBQoL-VN. The CVI was calculated after the second review. All I-CVI and S-CVI values for the 22 items were 1.0, indicating 100% expert agreement (Table 4).

**Table 3.** The 22-item CBQoL-VN.

| Domain | Item |
| --- | --- |
| **Health**<br>**and**<br>**lifestyle**<br>**(12 items)** | 1. Difficulty choosing or buying clothes |
|  | 2. Difficulty choosing groceries due to colour |
|  | 3. Difficulty knowing whether flowers, fruits, and food are still edible or spoiled due to colour |
|  | 4. Can't tell when food is cooked due to colour |
|  | 5. Not noticing change in colour of skin due to sunburn |
|  | 6. Not noticing change in the colour of mole on skin and wound (e.g. scratches) |
|  | 7. Not noticing blood in stools (faeces) |
|  | 8. Not noticing a change in colour of urine |
|  | 9. Being confused about colour of pills or other medication due to colour-coding |
|  | 10. Problems playing sports (e.g. colours of team clothing, colours of snooker balls etc) |
|  | 11. Difficulty reading or following maps (e.g. using electronic maps such as Google Maps or tracking locations for services like ride-hailing, motorbikes, or food delivery) |
|  | 12. Difficulty noticing and distinguishing traffic lights and traffic signs |
| **Emotions**<br>**(7 items)** | 13. Feeling anxious because of issues caused by problems seeing colours |
|  | 14. Feeling depressed because of issues caused by problems seeing colours |
|  | 15. Feeling unconfident because of issues caused by problems seeing colours |
|  | 16. Feeling embarrassed or humiliated because of colour blind issues |
|  | 17. Feeling different to other people because of issues caused by problems seeing colours |
|  | 18. Felt that had let down self or others due to problems seeing colours |
|  | 19. Avoiding conversations where colours are discussed |
| **Work and study**<br>**(3 items)** | 20. Difficulty performing work or other activities (e.g. analysing charts, identifying the colour of solutions or details etc) |
|  | 21. Being limited in choice of work or career |
|  | 22. Accomplishing less than would like at work or in career |

**Table 4.**
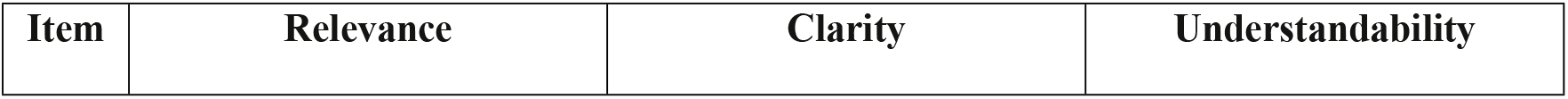
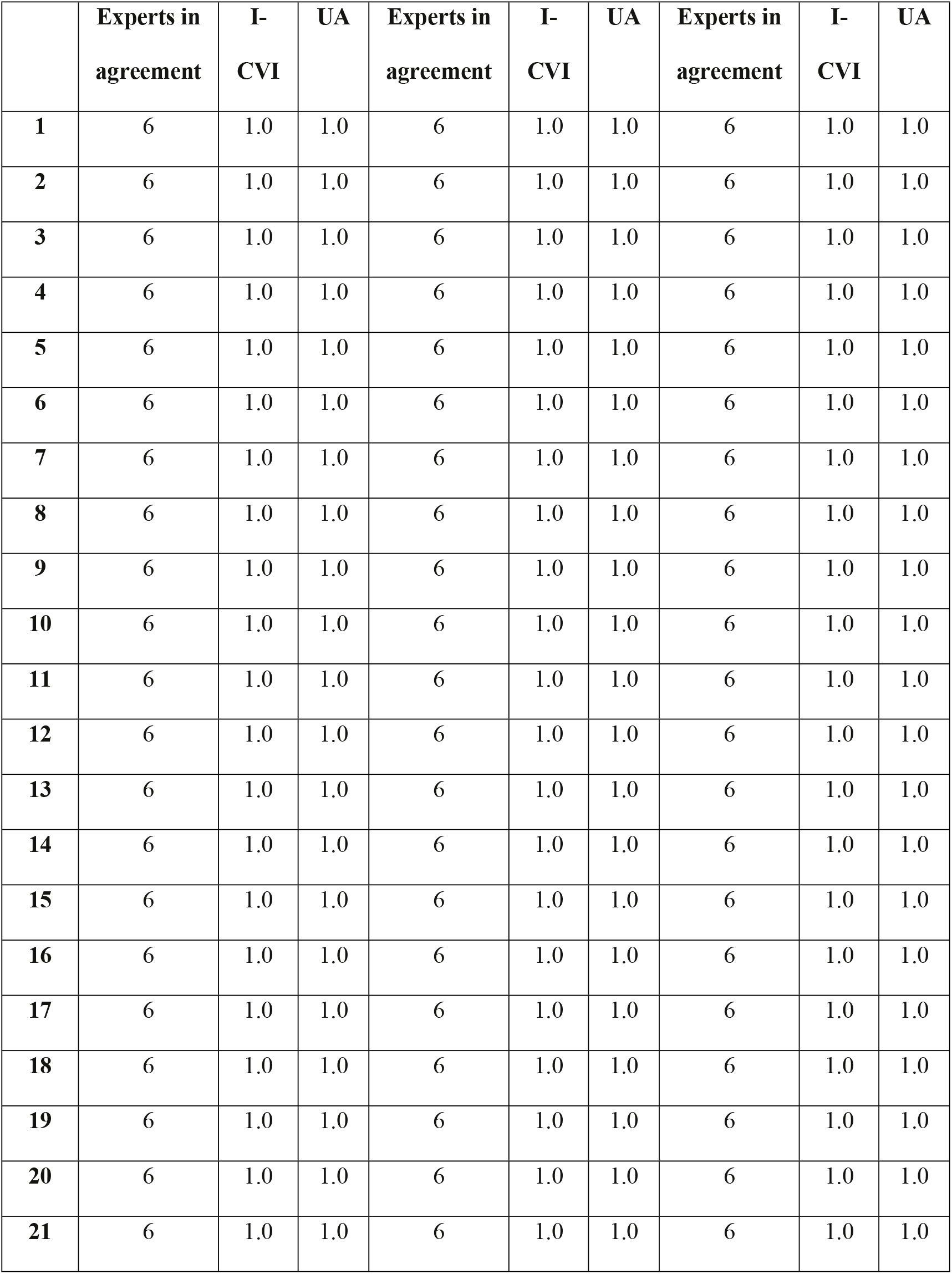
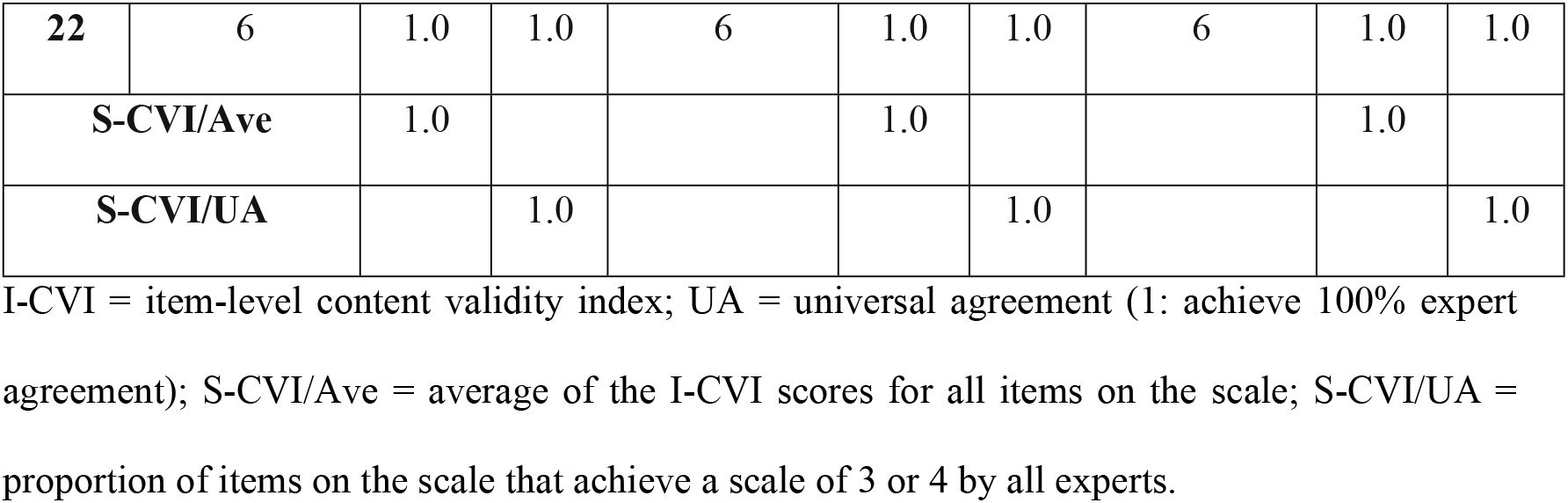
The CVI values for the 22-item CBQoL-VN by six experts.

| Item | Relevance | Clarity | Understandability |
| --- | --- | --- | --- |

|  | **Experts in**<br>**agreement** | **I-**<br>**CVI** | **UA** | **Experts in**<br>**agreement** | **I-**<br>**CVI** | **UA** | **Experts in**<br>**agreement** | **I-**<br>**CVI** | **UA** |
| --- | --- | --- | --- | --- | --- | --- | --- | --- | --- |
| **1** | 6 | 1.0 | 1.0 | 6 | 1.0 | 1.0 | 6 | 1.0 | 1.0 |
| **2** | 6 | 1.0 | 1.0 | 6 | 1.0 | 1.0 | 6 | 1.0 | 1.0 |
| **3** | 6 | 1.0 | 1.0 | 6 | 1.0 | 1.0 | 6 | 1.0 | 1.0 |
| **4** | 6 | 1.0 | 1.0 | 6 | 1.0 | 1.0 | 6 | 1.0 | 1.0 |
| **5** | 6 | 1.0 | 1.0 | 6 | 1.0 | 1.0 | 6 | 1.0 | 1.0 |
| **6** | 6 | 1.0 | 1.0 | 6 | 1.0 | 1.0 | 6 | 1.0 | 1.0 |
| **7** | 6 | 1.0 | 1.0 | 6 | 1.0 | 1.0 | 6 | 1.0 | 1.0 |
| **8** | 6 | 1.0 | 1.0 | 6 | 1.0 | 1.0 | 6 | 1.0 | 1.0 |
| **9** | 6 | 1.0 | 1.0 | 6 | 1.0 | 1.0 | 6 | 1.0 | 1.0 |
| **10** | 6 | 1.0 | 1.0 | 6 | 1.0 | 1.0 | 6 | 1.0 | 1.0 |
| **11** | 6 | 1.0 | 1.0 | 6 | 1.0 | 1.0 | 6 | 1.0 | 1.0 |
| **12** | 6 | 1.0 | 1.0 | 6 | 1.0 | 1.0 | 6 | 1.0 | 1.0 |
| **13** | 6 | 1.0 | 1.0 | 6 | 1.0 | 1.0 | 6 | 1.0 | 1.0 |
| **14** | 6 | 1.0 | 1.0 | 6 | 1.0 | 1.0 | 6 | 1.0 | 1.0 |
| **15** | 6 | 1.0 | 1.0 | 6 | 1.0 | 1.0 | 6 | 1.0 | 1.0 |
| **16** | 6 | 1.0 | 1.0 | 6 | 1.0 | 1.0 | 6 | 1.0 | 1.0 |
| **17** | 6 | 1.0 | 1.0 | 6 | 1.0 | 1.0 | 6 | 1.0 | 1.0 |
| **18** | 6 | 1.0 | 1.0 | 6 | 1.0 | 1.0 | 6 | 1.0 | 1.0 |
| **19** | 6 | 1.0 | 1.0 | 6 | 1.0 | 1.0 | 6 | 1.0 | 1.0 |
| **20** | 6 | 1.0 | 1.0 | 6 | 1.0 | 1.0 | 6 | 1.0 | 1.0 |
| **21** | 6 | 1.0 | 1.0 | 6 | 1.0 | 1.0 | 6 | 1.0 | 1.0 |
| 22 | 6 | 1.0 | 1.0 | 6 | 1.0 | 1.0 | 6 | 1.0 | 1.0 |
| **S-CVI/Ave** |  | 1.0 |  |  | 1.0 |  |  | 1.0 |  |
| **S-CVI/UA** |  |  | 1.0 |  |  | 1.0 |  |  | 1.0 |

I-CVI = item-level content validity index; UA = universal agreement (1: achieve 100% expert agreement); S-CVI/Ave = average of the I-CVI scores for all items on the scale; S-CVI/UA = proportion of items on the scale that achieve a scale of 3 or 4 by all experts.

### Reliability

Cronbach’s alpha for the 22-item CBQoL-VN was 0.95, indicating excellent internal consistency. By domain, Cronbach’s alpha values were 0.93 for “Health and lifestyle” (Items 1-12), 0.90 for “Emotions” (Items 13-19), and 0.89 for “Work and study” (Items 20-22), all of which demonstrated good reliability. Table 5 presents the corrected item-total correlations for the full scale and each domain. In all cases, each item was positively correlated with the total score of the remaining items.

**Table 5.** The corrected item-total correlations.

| Item | Corrected item-total correlation |  |  |  |
| --- | --- | --- | --- | --- |
|  | Full scale | Health and lifestyle | Emotions | Work and study |
| 1 | 0.51 | 0.59 | - | - |
| 2 | 0.54 | 0.62 | - | - |
| 3 | 0.73 | 0.80 | - | - |
| 4 | 0.63 | 0.70 | - | - |
| **5** | 0.52 | 0.69 | - | - |
| **6** | 0.48 | 0.63 | - | - |
| **7** | 0.77 | 0.84 | - | - |
| **8** | 0.76 | 0.84 | - | - |
| **9** | 0.82 | 0.67 | - | - |
| **10** | 0.76 | 0.68 | - | - |
| **11** | 0.62 | 0.49 | - | - |
| **12** | 0.73 | 0.68 | - | - |
| **13** | 0.69 | - | 0.58 | - |
| **14** | 0.74 | - | 0.85 | - |
| **15** | 0.68 | - | 0.85 | - |
| **16** | 0.68 | - | 0.69 | - |
| **17** | 0.37 | - | 0.42 | - |
| **18** | 0.73 | - | 0.82 | - |
| **19** | 0.78 | - | 0.74 | - |
| **20** | 0.80 | - | - | 0.82 |
| **21** | 0.73 | - | - | 0.81 |
| **22** | 0.68 | - | - | 0.71 |

### Discriminant validity

Table 6 presents the discriminant validity of the CBQoL-VN scores between participants with congenital CVD and those with normal colour vision. Participants with congenital CVD scored significantly lower across all CBQoL-VN domains compared with those with normal colour vision, supporting the discriminant validity of the instrument.

**Table 6.** Comparison of CBQoL-VN scores between congenital CVD and normal colour vision groups.

| CBQoL-VN<br>score | Congenital CVD<br>(n=30) |  | Normal colour vision<br>(n=30) |  | Significance<br>level |
| --- | --- | --- | --- | --- | --- |
| | Mean ± SD | Median<br>(IQR) | Mean ± SD | Median<br>(IQR) | |
| **Domain: Health**<br>**and lifestyle** | 5.13 ± 0.99 | 5.46<br>(1.35) | 5.94 ± 0.19 | 6.0<br>(0.02) | $p < 0.001$ |
| **Domain:**<br>**Emotions** | 5.01 ± 1.04 | 5.43<br>(1.82) | 5.96 ± 0.18 | 6.0 (0.0) | $p < 0.001$ |
| **Domain: Work**<br>**and study** | 4.89 ± 1.28 | 5.67<br>(2.08) | 5.97 ± 0.18 | 6.0 (0.0) | $p < 0.001$ |
| **Full scale** | 15.04 ±<br>3.01 | 16.60<br>(5.22) | 17.87 ±<br>0.55 | 18.0<br>(0.08) | $p < 0.001$ |

SD = standard deviation; IQR = interquartile range

## Discussion

The aims of this study were to translate and adapt the CBQoL into Vietnamese (CBQoL-VN) and to evaluate its validity and reliability in a Vietnamese congenital CVD population. The overall findings indicate a successful translation and cross-cultural adaptation, ensuring suitability for use in Vietnam.

The rigorous content validation process employed in this study ensured that the CBQoL-VN is not merely a linguistic translation, but a culturally and conceptually equivalent instrument for the Vietnamese population. The unanimous I-CVI and S-CVI values of 1.0 indicate that the expert panel considered all items highly relevant. During the review, several issues of redundancy and cultural adaptation were identified. The expert panel identified semantic overlaps in the emotional domain, specifically between items related to anxiety (Original CBQoL: Items 12 and 17) and self-esteem (Original CBQoL: Items 14 and 16). While these distinctions may be clear in the original English context, the nuances were obscured in the Vietnamese translation. Consequently, the removal of Items 16 and 17 aligns with best practices in questionnaire adaptation, which emphasize avoiding semantically similar items that could lead to participant confusion or response fatigue [16].

Furthermore, the review highlighted the necessity of modifications to achieve experiential equivalence. The distinction between “groceries” (Original CBQoL: Item 2) and “fruit” (Original CBQoL: Item 8) initially appeared ambiguous in the Vietnamese context. The revision of Item 8 (CBQoL-VN rearrangement: Item 3) to explicitly include “flowers, fruits, and food” ensures that the item captures a distinct aspect of daily living separate from general grocery shopping. This modification is consistent with guidelines by Beaton et al. (2000), which emphasize that cross-cultural adaptation often necessitates changing specific activities to reflect the target culture’s lifestyle while maintaining the original concept. The rephrasing of selected items (original CBQoL: Items 3, 9, and 22; CBQoL-VN rearrangement: Items 6, 11, and 20) further improved clarity and contextual relevance, particularly in reflecting Vietnamese daily lifestyle practices. Such adjustments strengthen the validity of the instrument and increase the likelihood that respondents will interpret items as intended.

Finally, the addition of a transportation-related item (CBQoL-VN: Item 12) to the health & lifestyle domain represents a critical contextual adaptation. While maintaining the structure of the original scale is important, guidelines on cross-cultural adaptation acknowledge that items may need to be added to capture phenomena unique to the new setting [17]. Given the complexity of traffic conditions in Vietnam, where mixed traffic and motorcycles are dominant [18], the ability to distinguish traffic signals and vehicle indicators is a crucial safety concern. The inclusion of this item ensures the CBQoL-VN accurately reflects the specific environmental challenges and safety risks faced by Vietnamese individuals with congenital CVD.

The internal consistency of the CBQoL-VN was found to be excellent, with a Cronbach’s alpha of 0.95 for the full scale. This indicates that the Vietnamese adaptation maintains the structural integrity of the initial instrument. Notably, this reliability is comparable to values reported in other recent adaptations, the Telugu version of the scale (Cronbach’s alpha = 0.70 to 0.90) [19]. The high alpha values across the individual domains of health and lifestyle, emotions, and work further confirm that the scale is a stable measure for diverse aspects of a patient’s life. While the original study by Barry et al. [7] established the three-factor structure of the instrument, they focused primarily on construct validity rather than reporting specific internal consistency coefficients for the final scale. Therefore, this study adds valuable psychometric data by confirming that these three domains possess high internal reliability within the Vietnamese population.

Discriminant validity was assessed by comparing the CBQoL-VN scores between individuals with congenital CVD and those with normal colour vision. The results revealed a statistically significant difference between the two groups, with the CVD group scoring significantly lower (indicating greater difficulty and lower quality of life) across the total scale and all subscales. This distinct separation confirms that the CBQoL-VN is specific to the impact of CVD and is not merely measuring general quality of life or generalized anxiety. These results align with findings from previous comparative studies [7,19], which consistently demonstrate that individuals with CVD face specific, measurable challenges in daily activities, occupational choices, and emotional well-being that are not present in the general population. Furthermore, research focusing exclusively on the quality of life within CVD cohorts has similarly reported reduced quality of life outcomes [5,6]. The ability of the CBQoL-VN to differentiate between these groups supports its utility as a clinical and research tool for identifying individuals who may require additional support or vocational counseling.

This study has several limitations. First, the small sample size of congenital CVD participants (n=30) precluded the use of factor analysis to statistically confirm the structural domains, although internal consistency remained high. This limitation is expected, given that congenital CVD has a relatively low prevalence globally, making it challenging to recruit large patient cohorts for psychometric analysis. Second, the questionnaire was specifically designed for individuals who are currently studying or employed, as they are better positioned to critically evaluate the impact of CVD on academic, occupational, and daily activities. This inclusion criterion further narrowed the recruitment base. Third, as Ishihara plates are designed to screen for protan and deutan defects, they are ineffective for detecting tritan-type deficiencies. Consequently, individuals with tritan defects may have been excluded from the sample.

## Conclusions

This study successfully translated and cross-culturally adapted the CBQoL into Vietnamese (CBQoL-VN). The adaptation process ensured cultural suitability, particularly through the refinement of items regarding daily activities and the inclusion of a transportation-specific item to reflect local traffic conditions. The scale also demonstrated strong discriminant validity, effectively distinguishing the specific quality-of-life burdens faced by individuals with congenital CVD compared to those with normal colour vision. The CBQoL-VN is a valid and reliable instrument for researchers and clinicians in Vietnam to assess the impact of colour vision deficiency, enabling more targeted vocational counseling and support.

## Data Availability

The relevant raw data used in this study is available with the DOI: https://doi.org/10.5281/zenodo.20154755.

## Acknowledgements

The authors wish to thank all the experts and participants for their contribution to this study.

